# Aerosol Jet Printing Enabled Dual-Function Electrochemical and Colorimetric Biosensor for SARS-CoV-2 Detection

**DOI:** 10.1101/2023.04.20.23288904

**Authors:** Li Liu, Zhiheng Xu, Adrian Moises Molina Vargas, Stephen J. Dollery, Michael G. Schrlau, Denis Cormier, Mitchell R. O’Connell, Gregory J. Tobin, Ke Du

## Abstract

An aerosol jet printing enabled dual-function biosensor for the sensitive detection of pathogens using SARS-CoV-2 RNA as an example has been developed. A CRISPR-Cas13: guide-RNA complex is activated in the presence of a target RNA, leading to the collateral trans-cleavage of ssRNA probes that contain a horseradish peroxidase (HRP) tag. This, in turn, catalyzes the oxidation of 3,3′,5,5′-tetramethylbenzidine (TMB) by HRP, resulting in a color change and electrochemical signal change. The colorimetric and electrochemical sensing protocol does not require complicated target amplification and probe immobilization and exhibits a detection sensitivity in the femtomolar range. Additionally, our biosensor demonstrates a wide dynamic range of 5 orders of magnitude. This low-cost aerosol inkjet printing technique allows for an amplification-free and integrated dual-function biosensor platform, which operates at physiological temperature and is designed for simple, rapid, and accurate point-of-care (POC) diagnostics in either low-resource settings or hospitals.

## INTRODUCTION

Respiratory tract infections caused by viruses post a significant challenge to the global public health.^1^ The unprecedented COVID-19 pandemic, for example, resulted in extremely high morbidity and mortality worldwide.^2^ While vaccines and antiviral drugs are the most effective specific strategies against pandemic viruses^3^, their development normally requires pre-clinical animal studies, clinical trials, and long production cycles, and thus they are generally not available at the early stages of a pandemic.^4^ On the other hand, accurate and timely diagnosis followed by isolation and treatment has been proved as an effective measure to slow down the transmission and reduce death rates. Quantitative Real-time reverse transcription polymerase chain reaction (qRT-PCR) with its superior sensitivity and specificity remains the gold standard for viral nucleic acid detection. However, operating a conventional qRT-PCR test is time-consuming, requiring specialized instruments and skilled personnel in a centralized lab which causes issues of distribution, affordability, and accessibility issues.^5^ Therefore, developing a rapid, simple, and highly sensitive diagnostic platform which can be analyzed at the point-of-care (POC) setting is crucial for the detection and control of future disease outbreaks.

Clustered regularly interspaced short palindromic repeats (CRISPR)-Cas based assays are an emerging technology for sensitive and specific detection of pathogenic DNA/RNA.^6–8^ A target-activated CRISPR-Cas complex enables collateral cleavage of single-stranded nucleic acid probes for signal amplification and detection of the intended target. Compared with conventional nucleic acid detection methods, CRISPR-based diagnostics have several advantages such as a high specificity due to the enzyme recognition of the pathogen-specific target sequences. Thus, false-positive signals can be eliminated to a great extent.^9^ In addition, CRISPR assays are typically performed at physiological temperatures without relying on sophisticated temperature control systems.^10^ CRISPR assays, especially using Cas13a, can complete a trans cleavage reaction within 10 min, which is ideal for rapid detection.^11^ In addition, CRISPR assays can be integrated with the established target amplification methods such as polymerase chain reaction (PCR),^12^ loop-mediated isothermal amplification (LAMP),^13^ and recombinase polymerase amplification (RPA)^14^ to further improve the detection sensitivity.

Moreover, CRISPR-Cas assays can work with various sensing mechanisms to achieve a multiplexing and amplification-free diagnostic platform. For example, electrochemical biosensors have drawn great interest due to their excellent sensitivity/specificity, cost-effectiveness, and ease of manufacturing.^15–17^ CRISPR-integrated electrochemical biosensors have been developed for biomarker detection of nucleic acids, proteins, and transcriptional regulation.^18–20^ However, most of these biosensors are fabricated by expensive microfabrication processes and require the immobilization of sensing probes on an electrode surface, which is complicated and time-consuming.^21,22^ Therefore, developing a new electrochemical biosensor platform that does not require probe immobilization is highly desired.

Here, we present a dual-function biosensor for the detection of infectious disease pathogens using SARS-CoV-2 as an example. This biosensor is an integrated three-electrode platform, fabricated by aerosol inkjet printing.^23,24^ Both colorimetric and electrochemical readouts are provided by the catalytic oxidation of 3,3′,5,5′-tetramethylbenzidine (TMB) with a horseradish peroxidase (HRP) tag. After activation of CRISPR-Cas13a by an RNA target and subsequent cleavage of the HRP labeled ssRNA probes, TMB substrate is added to produce colored oxidation product by a redox reaction. This reaction changes both the electrochemical properties and the color of the assay solution and is thus dual function. Without target amplification, a detection limit of 324 fM and 207 fM are achieved for colorimetric and electrochemical sensing, respectively. Both readouts demonstrate excellent linearity with target concentrations ranging from femtomolar to picomolar range and are highly specific against negative control samples. Thus, this dual function biosensor establishes a key technology that intended for POC detection of infectious diseases.

## EXPERIMENTAL SECTION

### Materials and reagents

MyOne™ Streptavidin C1 Dynabeads™ (10 mg/ml) and TMB Substrate Solution were purchased from Thermo Fisher Scientific, Inc. HRP Anti-Fluorescein antibody (1 mg/mL) was purchased from Abcam, Inc. Guide RNA and RNA reporters were purchased from IDT. Inc. Lbu-Cas13a protein was prepared based on our established protocols, described in our previous publication.^25^

### Preparation of target RNA segments

The viral genetic sequences of SARS-CoV-2 (positive sample, 703 nucleotides), SARS-1 (negative control, 660 nucleotides), and Influenza A (negative, control, 547 nucleotides) were amplified from plasmids pUC57-SARS-CoV-2, pUC57-SARS-CoV-1, and p1041-Influenza A via PCR and TA cloned into vector TOPO-TA, directly following a T7 promoter (Invitrogen, Inc). To generate linear template DNA for *in vitro* transcription, plasmids were digested with *Hind*III (New England Biolabs, Inc.), EtOH precipitated, and resuspended in dH_2_O. RNA fragments were synthesized by T7 runoff reactions with linear plasmid DNA as a template using the RiboMAX large scale production system (Promega, Inc.) at 37°C for 4 hrs. Following RNA synthesis, the templates were degraded using DNaseI. Proteins, template fragments, and excess nucleotides were removed by silica gel membrane column purification (RNeasy-QIAGEN, Inc). RNA size and preliminary quantification were performed by agarose-TBE gel electrophoresis with a RiboRuler RNA ladder (ThermoFisher, Inc). Quantification was verified by UV absorption spectroscopy at 260 nm. All the target sequences are available in the supporting information (**Table S2**).

### CRISPR Lbu-Cas13a trans-cleavage

CRISPR Cas13a/guide RNA complexes were formed by incubating 1 μM of CRISPR Lbu-Cas13a and 1.1 μM of gRNA in 6 μL of 5× Standard Buffer (250 mM KCl, 100 mM HEPES, 25 mM MgCl_2_, 5 mM DTT, 25% Glycerol, pH 6.8) in 24 μL of RNase-free water at 37°C for 2 min. The reaction was then allowed to incubate at room temperature for another 8 min and placed on ice for later use. For each reaction, we mixed 4 μL of Cas13a/gRNA complex, 20 μL of RNase-free water, 8 μL of 5× Standard Buffer, 4 μL of target RNA, and 4 μL of fluorescein-biotin ssRNA (10 μM). The complex solution was incubated at 37°C for 15 min.

### Conjugating the RNA reporters onto the HRP Anti-Fluorescein antibodies

After the trans-cleavage of the ssRNA reporters, 20 μL of the HRP Anti-Fluorescein antibodies (0.6 μg/ml) were added into the mixture and incubated on a rotary mixer at room temperature for 30 min.

### Magnetic bead isolation

The streptavidin-coated magnetic beads were washed three times with a washing buffer (1×PBS containing 0.01% Tween 20). Then, 60 μL of the CRISPR Cas13a-HRP complex solution was added to 20 μL of beads solution. The mixture was incubated on a rotary mixer at room temperature for 30 min. The HRP-magnetic beads were separated from the unbound ssRNA reporters by magnetic separation.

### Colorimetric detection

The magnetic bead solution was then washed with washing buffer three times. Afterward, 20 μL of the TMB substrate solution was added into the magnetic bead solution. The colorimetric changes were visually observed after a 15 min reaction. The absorption spectrum was recorded with a microplate reader (Spectramax iD3, Molecular Devices, USA) by removing the supernatant from the beads solution and diluting to 100 μL in nuclease-free water.

### RGB value analysis

Images were taken by a smartphone camera (iPhone 12) and the changes in RGB values for each reaction were extracted for analysis. A 500-pixel x 500-pixel square in each reaction pool was extracted (n=5) and analyzed by ImageJ, where the color RGB value was recorded.

### Fabrication of the graphene electrode

The 3-electrode platform includes a working electrode, a counter electrode, and a reference electrode. An aerosol jet (Aerosol Jet 300, Optomec Inc. US) with aerodynamic focusing was used to precisely deposit graphene ink (Sigma Inc. US) onto the polyethylene terephthalate (PET)-based printed electronics substrate (Novacentrix, Novele™ IJ-220). The ink was placed in an atomizer that produces ink droplets with a size between 1 and 5 μm. The aerosol mist was then delivered to the deposition head, which was focused by the sheath gas that acts as an annulus around the aerosol. As the sheath gas and aerosol passed through the nozzle, they were accelerated, and the aerosol was “focused” into a tight stream of droplets that flows within the sheath gas. The resulting high-speed particle stream remained focused over a distance of 2 mm from the nozzle to the substrate without compromising the resolution. Once the chip was fabricated, photonic curing was used to rapidly and efficiently convert the graphene ink solution into high-quality thin-film electrodes.^26^ Samples were passed through a Novacentrix Pulseforge 3300 machine to expose graphene to the incident energy. The PulseForge 3300 tool is capable of delivering peak power up to 100 kW/cm^2^ in pulses as short as 30 ms. In this work, each sample was exposed to two 200 V pulses with a pulse frequency of 2.8 Hz and a duration of 1,200 ms.

### Characterization of the 3-electrode testing chip

The electrode morphology was scanned using a profilometer (Modular Standard Optical Profilometer, Nanovea ST400). The dimensions of the three-electrode section and the thickness of various materials were determined by the profilometer. The stability and reproductivity for the 3-electrode system were also tested. Twenty microliters of H_2_SO_4_ background electrolyte (0.05 M) were added into the reservoir. Cyclic voltammetry was set between 0 and 1.2 V/ The speed rate was 0.1 V/s and the number of scans was five cycles.

### Electrochemical detection

After the magnetic bead isolation, 20 μL of the reacted TMB solution was removed from the tube and placed into the electrochemical platform reservoir. The chronoamperometric response was obtained at + 3mV over 60 s on a potentiostat (Reference 600™, Gamry Instruments Inc, USA) with a graphene counter electrode and a reference electrode.

## RESULTS

The working principle of the dual-function biosensor is illustrated in **Figure 1A**. In this protocol, an RNA fragment of the S gene of SARS-CoV-2 is detected by the LbuCas13a:crRNA complex. In the presence of the target gene, the dual labeled fluorescein and biotin ssRNA reporters are cleaved by the CRISPR-Cas13a complex. In the next step, the HRP anti-fluorescein antibody is added to the solution, which can be conjugated with the fluorescein on the reporter side of the ssRNA. After magnetic separation, the conjugates are present on the magnetic beads while the unbounded HRP anti-fluorescein antibody is removed from the supernatant. In the last step, TMB substrate solution is added to the washed magnetic beads, allowing the HRP on the magnetic beads to catalyze TMB to the blue product TMB^2+^. For electrochemical detection, the amount of HRP attached to the magnetic beads varies with the target RNA concentration due to the amount of trans-cleaved probes. HRP reacts with TMB for signal amplification and current detection. For colorimetric detection, the HRP conjugated on the magnetic beads can catalyze the oxidation reaction of TMB. This reaction presents a color change from colorless to blue and can be detected by a microplate reader. The inset in **Figure 1A** shows the details of the activated and inactivated Cas13a reaction. When the Cas13 enzyme is activated, no HRP remains on the beads, thus TMB cannot be oxidized. On the other hand, inactivated Cas13 enzyme does not cleave the reporter probes, leaving HRP on the magnetic beads and oxidizing TMB, which increases H_2_O in the solution and decreases the current. **Figure 1B** shows the segment of the target and the guide RNA sequences. We used the gold electrodes to monitor SARS-CoV-2 RNA-activated LbuCas13a:crRNA complex endonuclease activity. As shown in **Figure 1C**, the current signal increases significantly from 0.30 to 0.84 after the addition of the LbuCas13a:crRNA complex, indicating that the CRISPR complex is active.

**Figure 1.**
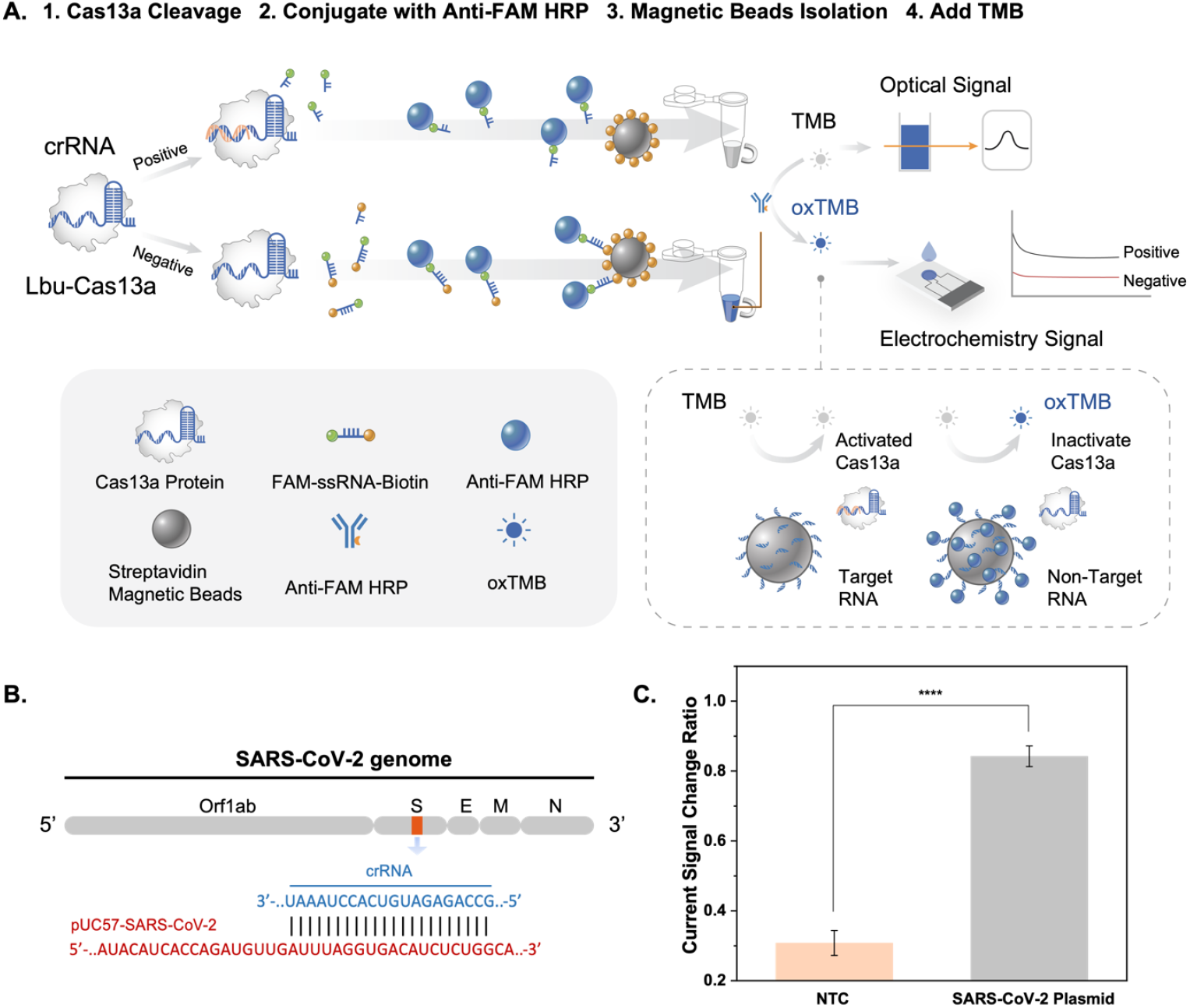
A. Schematic of the reaction strategy: Step 1. CRISPR-Cas13a:crRNA complex cleaves an ssRNA probe; Step 2. Anti-FAM HRP binds the FAM labeled ssRNA probe; Step 3. The non-cleaved biotin-labeled ssRNA probe is isolated by streptavidin magnetic beads; Step 4. Colorimetric reaction and current detection with TMB oxidization. B. SARS-CoV-2 gene location, target site, and guide RNA sequence. C. Current Signal Change Ratio with and without SARS-CoV-2 target, showing ∼3-fold difference in signal. The statistical significances are calculated by *t*-test (****P ≤ 0.0001). Error bars: SD, n=3.

The fabrication process of the chip (10 mm × 6 mm) is shown in **Figure 2A**. The surface topography was characterized using optical profilometry (**Figure 2B**) and cyclic voltammetry (**Figure 2C**). The thickness of the graphene electrode is 3 ± 0.2 μm, showing a very smooth surface (**Figure S1**). The repeatability and reproducibility of the gold electrodes were tested by tracking their response to H_2_SO_4_ solutions over a set period. The electrode substrate was stored in a closed container and the response to the H_2_SO_4_ solution (0.05 mM) was recorded. **Figure 2C** shows several representative voltammetry curves of the graphene electrodes. In the voltammograms of the three measurements performed on the same day (lines 1, 2, and 3), 98.0% of the original peaks and potentials were found to be retained, demonstrating the high stability of the chip. Similarly, the response of the H_2_SO_4_ solution (0.05 mM) was recorded on different days (one day apart, lines 4, 5, 6). Voltammograms show similar trends with the same current magnitude at the desired potential with an RSD of 5.81%, demonstrating the high repeatability and reproducibility of the working electrode.^27^

**Figure 2.**
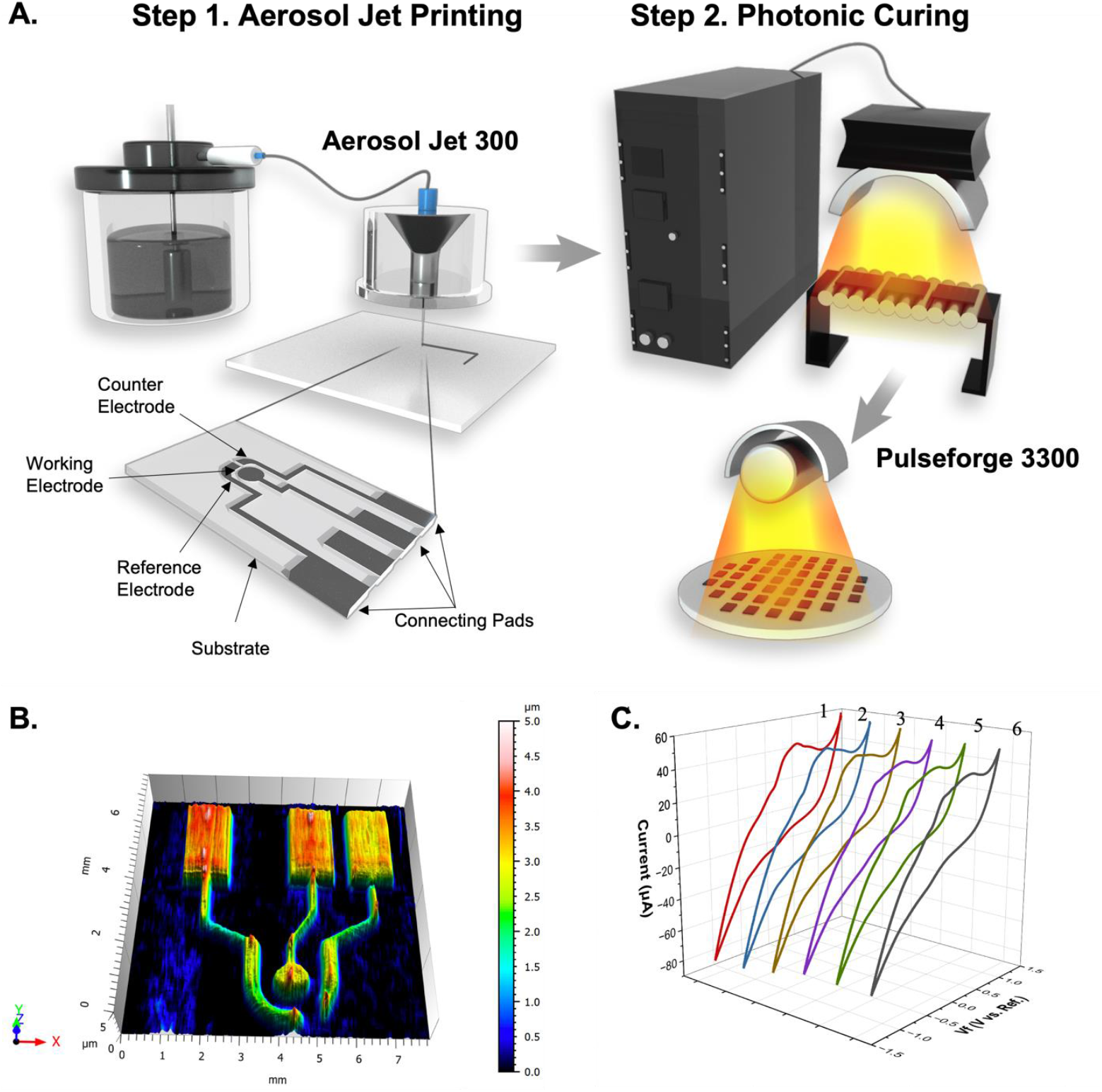
A. (1) Schematic of the aerosol jet printing process for the dual-function biosensor and the graphene electrode with a working electrode, a counter electrode, and a reference electrode. (2) The photonic curing system for the graphene electrode. B. Profilometer scan of the graphene electrode, showing a smooth surface with only ± 0.2 μm variation. C. Graphene electrode voltammetry curves for H_2_SO_4_ solution (0.05 mM). Lines 1, 2, and 3 are scans on the same day, and lines 4, 5, and 6 are scans from different days (one day apart).

In our assay, increasing concentrations of target RNA in the sample results in more HRP released from the magnetic beads, leaving less HRP enzyme to oxidize TMB. As shown in **Figure 3**, the absorbance of the characteristic peak at 650 nm derives from the catalyzed reaction product of TMB, indicating that HRP attached to the magnetic beads possesses good catalytic ability. As expected, the UV-vis absorption peak gradually decreases with the increasing target RNA concentration from 100 fM to 1 nM (**Figure 3A**). **Figure 3B** shows the absorption measurements for the different negative controls (SARS-1, Influenza A, and no target), which are noticeably different from the positive samples. In **Figure 3C**, the absorbance is linearly correlated with the change of target RNA concentration from 100 fM to 1 nM (R^2^ = 0.91385). Therefore, the detection limit was determined to be 324.1 fM based on the 3σ rule.^28^ The inset of **Figure 3C** is a photograph of the reaction solution in daylight associated with the increase of target RNA concentration. We note that the color of the reaction mixture changes continuously from dark to light with increasing target concentration, which can be identified by naked eye. The peak absorption of the positive and negative samples is shown in **Figure 3D** and the inset is a photograph of the sample, clearly showing the difference.

**Figure 3.**
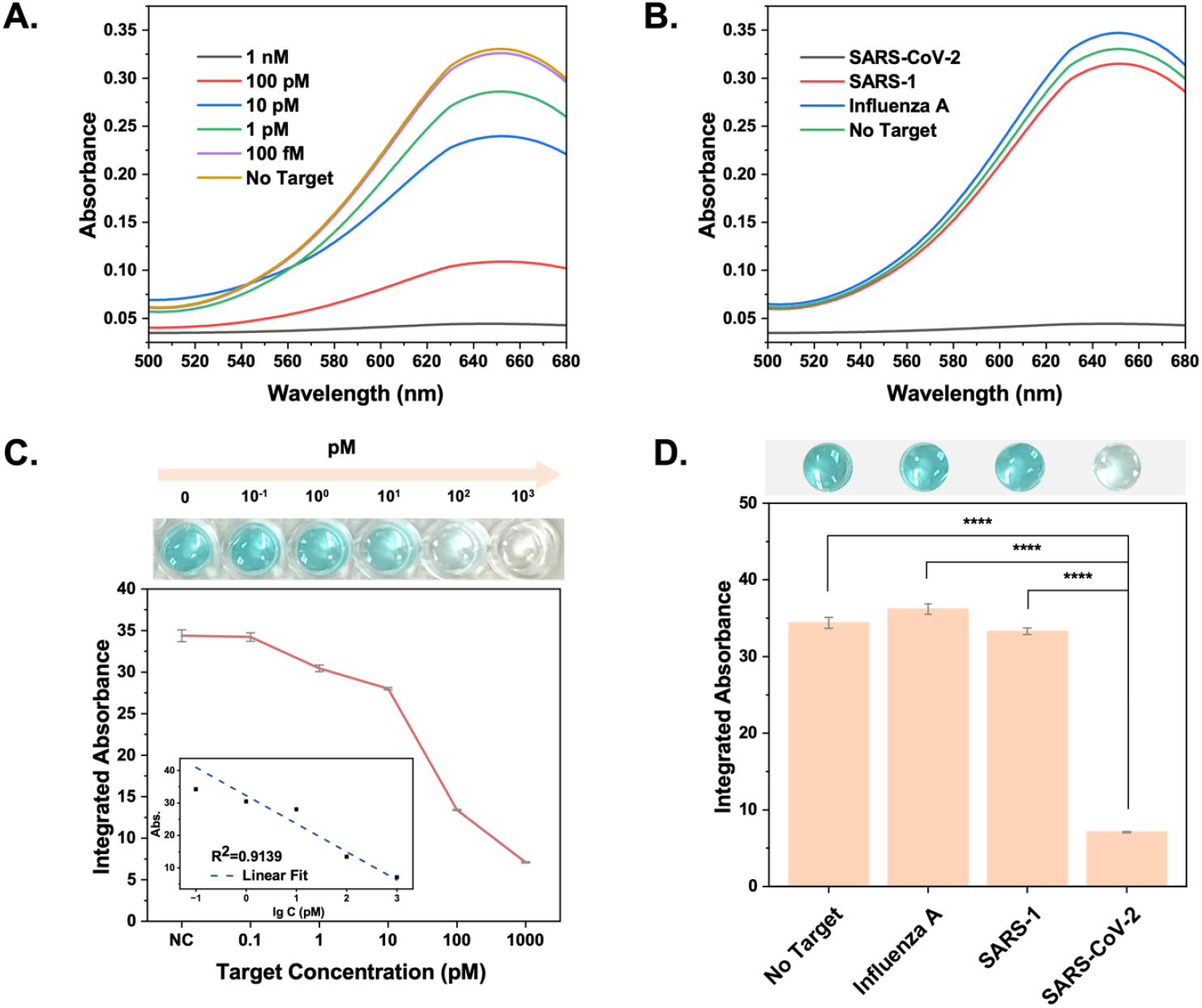
A. UV-vis spectrum vs. SARS-CoV-2 RNA concentration after RNA detection by our assay. B. UV-vis spectrum vs. positive and negative target. C. Top: Photographs of the test solution with different SARS-CoV-2 concentrations under daylight. Bottom: The relationship between the integrated absorbance and the concentration of SARS-CoV-2 RNA. Inset: The calibration curve for the integrated absorbance value vs. The logarithm of the target concentration. Error bars denote standard deviation (n = 5). D. Top: Photographs of the test solution of different targets under daylight. Detection specificity of the SARS-CoV-2 RNA target compared with other targets. Error bars denote standard deviation (n = 5). The asterisks represent statistical significance according to a *t*-test of *P ≤ 0.05, **P ≤ 0.01, ***P ≤ 0.001, ****P ≤ 0.0001.

After optimizing reaction conditions, the analytical performance of our chip was investigated. As shown in **Figure 4A**, the color of the substrate changes from blue to colorless as the concentration of SARS-CoV-2 RNA increases. By analyzing the RGB values, we found that the R values are linearly correlated with target RNA concentration ranging from 100 fM to 1 nM (R^2^ = 0.95081). (**Table S1**). Therefore, the detection limit was determined to be 194.7 fM according to the 3σ rule. The specificity of our chip was also investigated by using SARS-1 and Influenza A as negative samples. As shown in **Figure 4B**, without target input, the solution shows no significant color change, while the addition of target RNA triggers a dramatic color change. Similar to the no target sample, this color change can easily be observed by naked eye. This result indicates that our strategy can be used for the detection of SARS-CoV-2 RNA with a high sensitivity and specificity.

**Figure 4.**
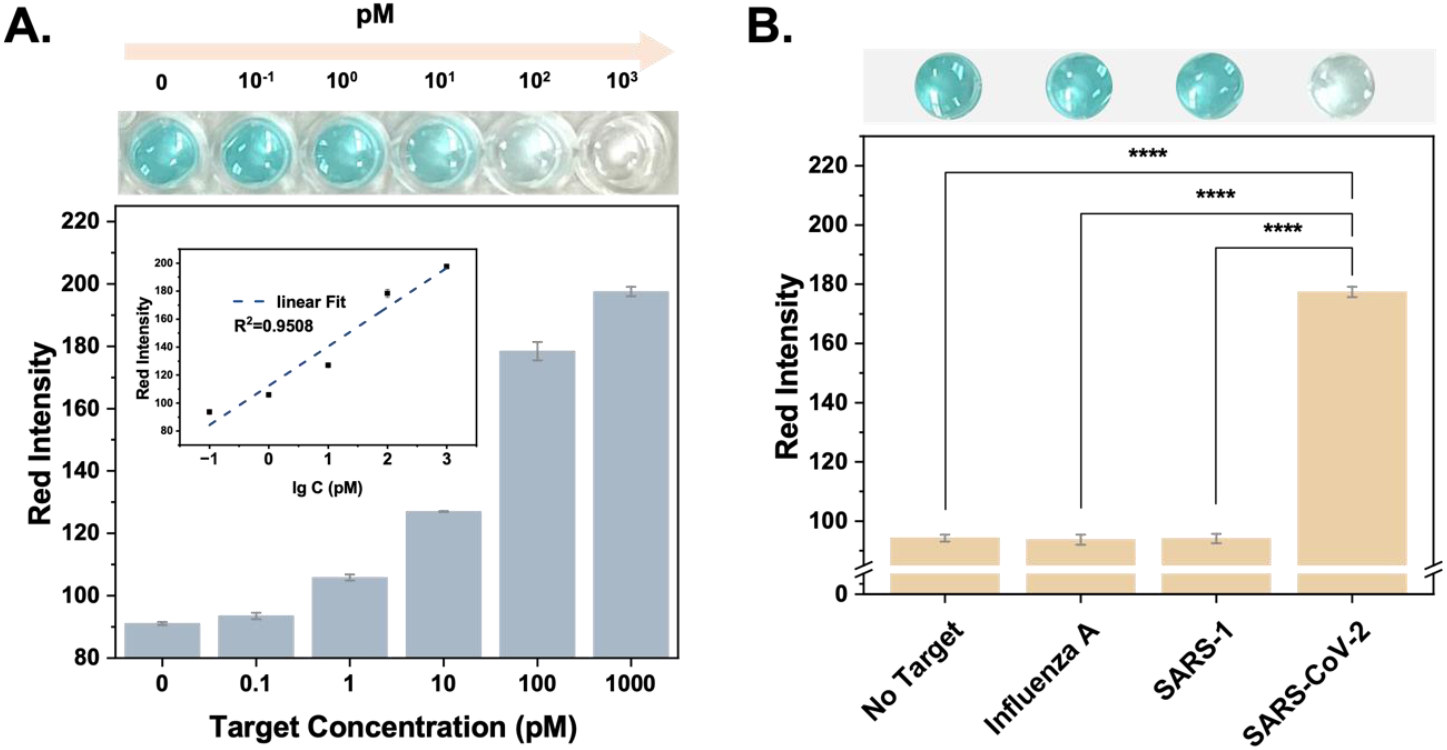
A. Top: Photographs of the test solution with different SARS-CoV-2 RNA concentrations under daylight. Bottom: The relationship between the R value and the concentration of the SARS-CoV-2 RNA. Inset: Calibration curve of the R value vs. The logarithm of the target concentration. Error bars denote standard deviation (n = 5). B. Top: Photographs of the test solution with different RNA targets under daylight. Bottom: Detection specificity of the SARS-CoV-2 RNA target compared with other targets. Error bars denote standard deviation (n = 5). The asterisks represent statistical significance according to a *t*-test. *P ≤ 0.05, **P ≤ 0.01, ***P ≤ 0.001, ****P ≤ 0.0001.

Electrochemical measurements were also performed to quantitatively assess the detection limit and dynamic range of the target. In the TMB substrate solution, the chronoamperometric response at +3 mV increases with increasing target RNA concentration (**Figure 5A**). A linear relationship was observed between chronoamperometric response and target RNA concentration ranging from 100 fM to 10 nM with a correlation coefficient (**Figure 5B**). The detection limit was determined to be 163 fM based on the 3σ rule. **Figure 5C** and **Figure 5D** show the comparison of the current curves and different current values for chronoamperometric reactions for positive and negative targets, showing clear differences between positive and negative groups. A comparison of the linear fits of R values and current values for different concentrations of SARS-CoV-2 targets is shown in **Figure 6**. Both readings show excellent linearity over the range of target concentration from femtomolar to picomolar, confirming the high sensitivity and reliability of the biosensor.

**Figure 5.**
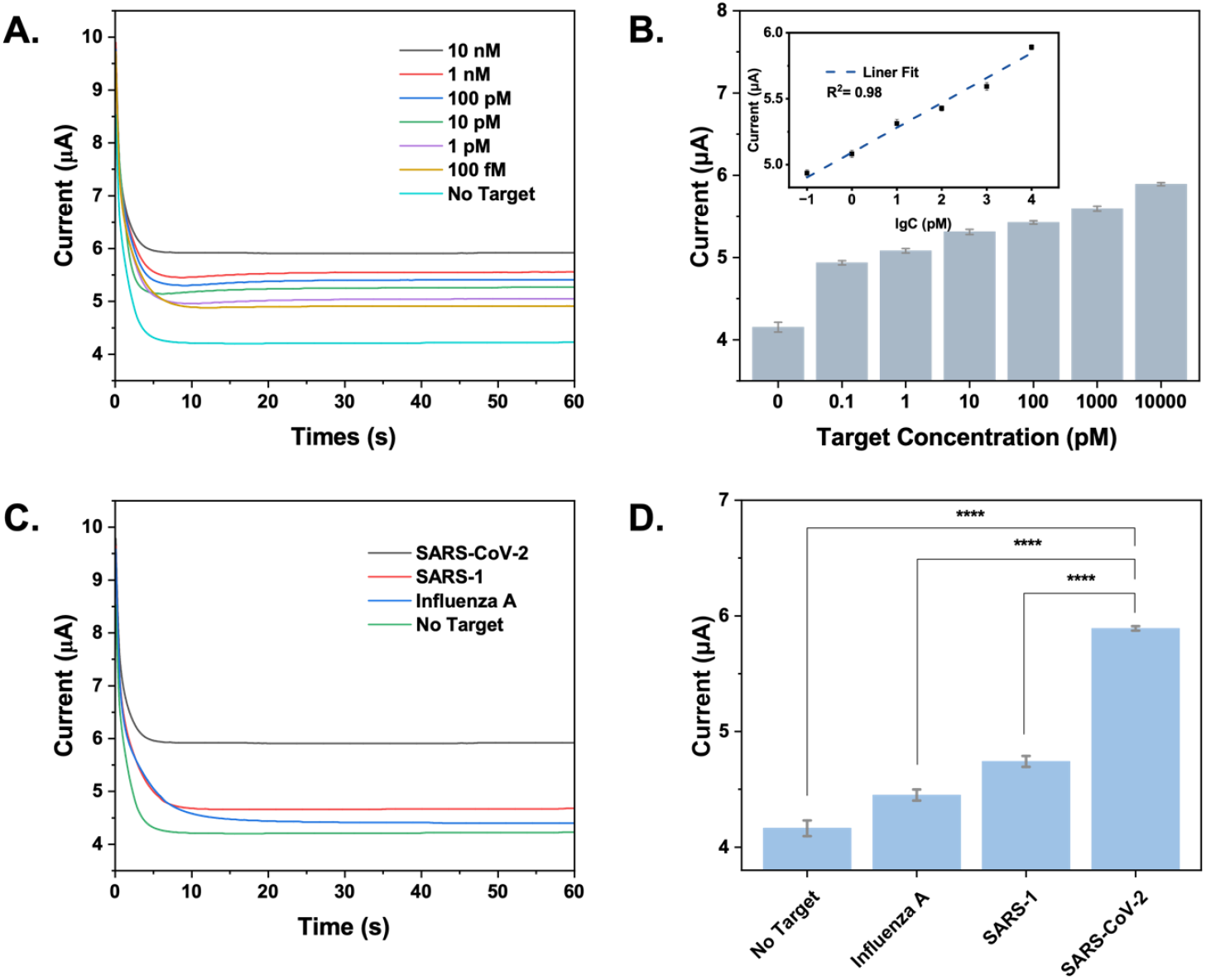
A. Chronoamperometric responses of SARS-CoV-2 RNA targets at different concentrations. B. Plot of current of last 30s mean value versus different concentrations of SARS-CoV-2 target. Error bars denote standard deviation (n = 5). Inset: Calibration curve of the current vs. the logarithm of the target concentration. C. Chronoamperometric responses of the positive and negative targets. D. Detection specificity of the SARS-CoV-2 RNA target compared with other targets. Error bars denote standard deviation (n = 5). The asterisks represent statistical significance according to a *t*-test. *P ≤ 0.05, **P ≤ 0.01, ***P ≤ 0.001, ****P ≤ 0.0001.

**Figure 6.**
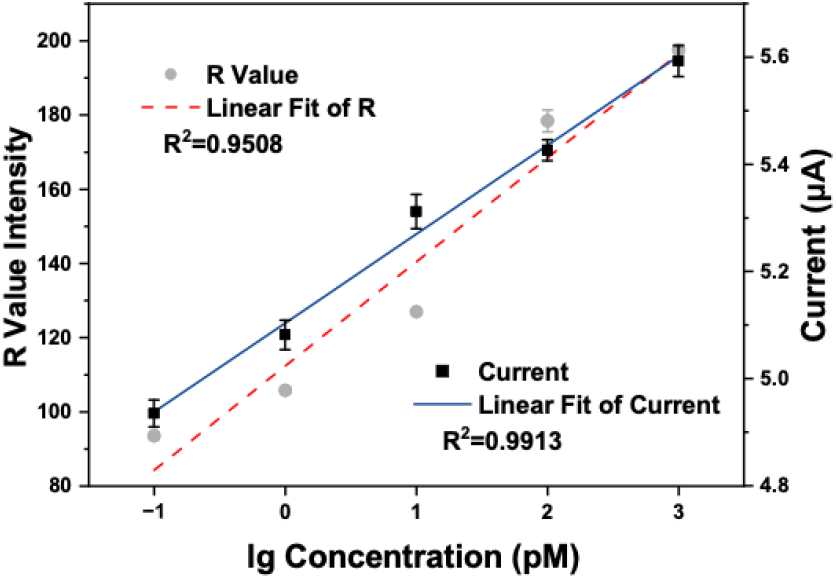
R value intensity and measured current vs. target RNA concentration with a R^2^ value of 0.9508 and 0.9913, respectively. Error bars denote standard deviation (n = 5).

## DISCUSSION

Our reaction chemistry is highly specific and reliable. First, specific guide RNA sequences only recognize specific target RNAs due to the characteristic specificity of the CRISPR guide RNA sensing the target sequence, making it more specific than other sensing methods, such as immunoassays.^7^ We show that we can differentiate SARS-CoV-2 from negative controls such as Influenza and SARS-CoV-1 at various concentrations. Second, in our reaction, the signal indicator Anti-FAM HRP is conjugated on the magnetic beads via ssRNA linkers. After the washing step, the impurities in the reaction are washed away thus leaving the final reactants with a relatively pure HRP enzyme to react with the TMB substrate. The high purity of the analyte is critical for electrochemical detection, as the presence of impurities in the reaction solution can cause current changes and reduce the detection sensitivity.^29^ Third, our amplification-free detection method simplifies the whole reaction and is designed for POC applications. In addition, a useful biosensor should have good stability during the sensing process. By observing the response of our biosensor exposed to sulfuric acid (0.05 mM) over a set time for 12 cycles, voltammograms of multiple measurements on the same chip were compared, showing a very high retention of the original peaks and potentials.

One unique advantage of our biosensor is the dual-function detection scheme as we can track both electrochemical signal change and colorimetric signal change simultaneously. Reading color changes with the naked eye is simple and straightforward, which is desired for rapid POC applications.^30^ On the other hand, the electrochemical sensing provides a more quantitative readout, which is crucial to monitor the viral load in the sample when necessary. Importantly, a similar detection limit for colorimetric and electrochemical sensing is achieved. This dual-function platform reduces the false-positive and false-negative results caused by the variation of target samples and test chips.

Our electrochemical chip was fabricated by aerosol-jet printing, which is highly stable and inexpensive. Aerosol-jet printing technology is a process of depositing extremely fine droplets (2-5 μm diameter) of ink suspended in an aerosol onto a substrate to form patterns. The aerosol used in this process acts as a carrier to deliver the ink to the surface, providing a more consistent and controlled deposition than traditional liquid-based inkjet technologies.^24^ One of the main advantages of aerosol-jet printing technology is its ability to produce high-resolution patterns with precise control over the size and distribution of the ink droplets. This high level of control enables the creation of intricate and detailed designs, making it ideal for a wide range of applications, including printing on flexible and transparent substrates.^31^ Another advantage of aerosol-jet printing technology is its ability to print on a wide range of materials, including those that are difficult to print on using traditional liquid-based inkjet technologies. This is due to the unique properties of aerosol, which allow it to conform to irregular surfaces and maintain its stability, even on porous or uneven substrates.^32,33^

Our biosensor is based on a highly specific CRISPR-Cas13 assay and a highly sensitive electrochemical detection platform. Without target amplification, a detection limit of 200 fM is achieved, which is 500 time more sensitive than our previous results of using pure liquid phase CRISPR-Ca13a reactions.^34^ This dual function of electrochemical and colorimetric approach also avoids photobleaching problems in fluorescence based sensing. In the future, our assay can be combined with RT-LAMP or RT-RPA methods to extend the detection limit by several orders of magnitude.^35–37^ In addition, by simply changing the design of guide RNA sequences, our assay can be used to detect a wide range of infections and diseases, such as cancers and sepsis, which are the main causes of premature deaths.^38^

## Data Availability

All data produced in the present study are available upon reasonable request to the authors

## Acknowledgements

This work was supported by the National Institute of General Medical Sciences of the National Institutes of Health under Award Number R35GM142763 (to K.D.), and R35GM133462 (to M.R.O.).

